# Pilot Evaluation of Patient-Assisted Motorized Transvaginal Ultrasound Acquisition During Ovarian Stimulation Monitoring

**DOI:** 10.64898/2026.06.21.26356181

**Authors:** Tal Shavit, Pietro Bortoletto, Johanna Szychter, Sari Mendel, Yael Corcos, John Petrozza, Nadia Prisant

## Abstract

This prospective first-in-human pilot study evaluated the feasibility, safety, and preliminary clinical performance of patient-assisted motorized transvaginal ultrasound during ovarian stimulation monitoring. Ten women underwent 19 investigational acquisitions following standard-of-care ultrasound. Participants inserted and stabilized the probe, while a predefined motorized rotational sequence was operated under clinical supervision. Eighteen of 19 acquisitions (94.7%) generated evaluable imaging datasets, with bilateral ovarian visualization in 16 of 18 (88.9%). No adverse events were observed, although three non-serious technical device deficiencies occurred. Investigational follicle counts ≥14 mm correlated with mature oocyte recovery (ρ=0.83), and endometrial thickness categories agreed in 14 of 15 paired examinations (93%). All participants expressed willingness to reuse the system. These findings support the feasibility of supervised patient-assisted acquisition and warrant larger studies to assess reproducibility and clinical performance.

## Objective

Transvaginal ultrasound is central to ovarian stimulation monitoring but requires repeated examinations by trained operators. Prior studies of self-operated endovaginal telemonitoring (SOET) and home vaginal ultrasound have shown that selected patients can participate in image acquisition, although these approaches relied on active patient navigation and real-time remote guidance (1,2). We prospectively evaluated a different acquisition model in which participants inserted and stabilized the transvaginal probe while images were acquired through a predefined motorized rotational sequence operated by a supervising healthcare professional.

## Study Design

Ten women undergoing ovarian stimulation for in vitro fertilization or fertility preservation were enrolled at a single IVF center. Each investigational examination immediately followed standard transvaginal ultrasound performed by one of two experienced sonographers using a Voluson S10 Expert system. The investigational prototype incorporated a transvaginal probe with an external motorized rotational mechanism. Participants inserted and stabilized the transvaginal probe. Motorized rotation was generated by an external pulley system that rotated the distal portion of the probe from −180° to +180° around its longitudinal axis according to predefined angular steps. The rotational sequence was predetermined and was not modified according to the ultrasound images displayed during acquisition.

Participants received no formal sonographic training and no real-time anatomical coaching. Investigational images were not used for treatment decisions. Feasibility was defined as generation of an evaluable imaging dataset. Secondary exploratory outcomes included bilateral ovarian visualization, safety, follicular assessment, endometrial thickness, and patient-reported experience. Follicular analyses included only ovaries visualized by both modalities. Individual follicles were not matched between sequential examinations; follicular size comparisons were therefore performed at the ovary level using the median diameter of follicles ≥10 mm. Counts of follicles ≥14 mm on the final pre-trigger examination were correlated with mature oocyte recovery (3). Analyses were descriptive and unadjusted.

## Results

Nineteen investigational acquisitions were attempted. Eighteen (94.7%) generated evaluable datasets, and bilateral ovarian visualization was achieved in 16 of 18 evaluable examinations (88.9%); 2 examinations had partial ovarian visualization. No adverse events, adverse device effects, vaginal injury, bleeding, or infection were observed. Three prototype-related technical device deficiencies occurred: two involved increased rotational resistance related to device setup, and one involved fracture of an external rotation handle during transportation. All three affected examinations were completed, none resulted in participant harm or clinical consequences, and neither issue recurred following corrective actions.

Individual follicles were not matched one-to-one between examinations; follicular size was therefore compared at the ovary level. Among 23 ovaries with measurable follicles ≥10 mm in both modalities, the median absolute difference between ovary-level median follicular diameters was 2.0 mm (IQR 0.5–3.75 mm). These exploratory measurements were not considered sufficient to establish interchangeability between modalities, consistent with known measurement variability in follicular ultrasound assessment (4,5). However, counts of follicles ≥14 mm correlated strongly with mature oocyte recovery for both investigational and standard ultrasound (Spearman ρ=0.83, p=0.01 and ρ=0.80, p=0.02, respectively). Endometrial thickness measurements were available in 15 paired examinations. The mean signed difference between investigational and standard ultrasound was −0.22 mm, the median absolute difference was 0.70 mm, and the maximum absolute difference was 1.75 mm. Agreement for endometrial thickness ≤7 mm versus >7 mm was 93% (14/15; Cohen κ=0.86). However, given the small number of paired observations and only one discordant classification, the kappa estimate is imprecise.

Patient-reported experience was also favorable. All participants reported willingness to use the system again, and overall examination experience was significantly better with investigational acquisition than with standard transvaginal ultrasound (Wilcoxon signed-rank p=0.002).

## Conclusions

These findings support the feasibility of supervised patient-assisted transvaginal ultrasound acquisition using controlled motorized rotation, with no safety events observed in this pilot study. However, the small number of acquisitions does not allow uncommon adverse events to be excluded, and the occurrence of three prototype-related technical deficiencies underscores the need for continued evaluation of device refinement and reliability during development.

The results do not establish interchangeability with standard transvaginal ultrasound, and investigational imaging was not used for clinical decisions. Nevertheless, the observed association between investigational follicle counts ≥14 mm and mature oocyte recovery, together with favorable patient-reported experience, supports larger prospective evaluation. Future studies should assess reproducibility and formal agreement against standard transvaginal ultrasound and determine whether progressively less supervised acquisition can support clinical decision-making.

**Table 1.**
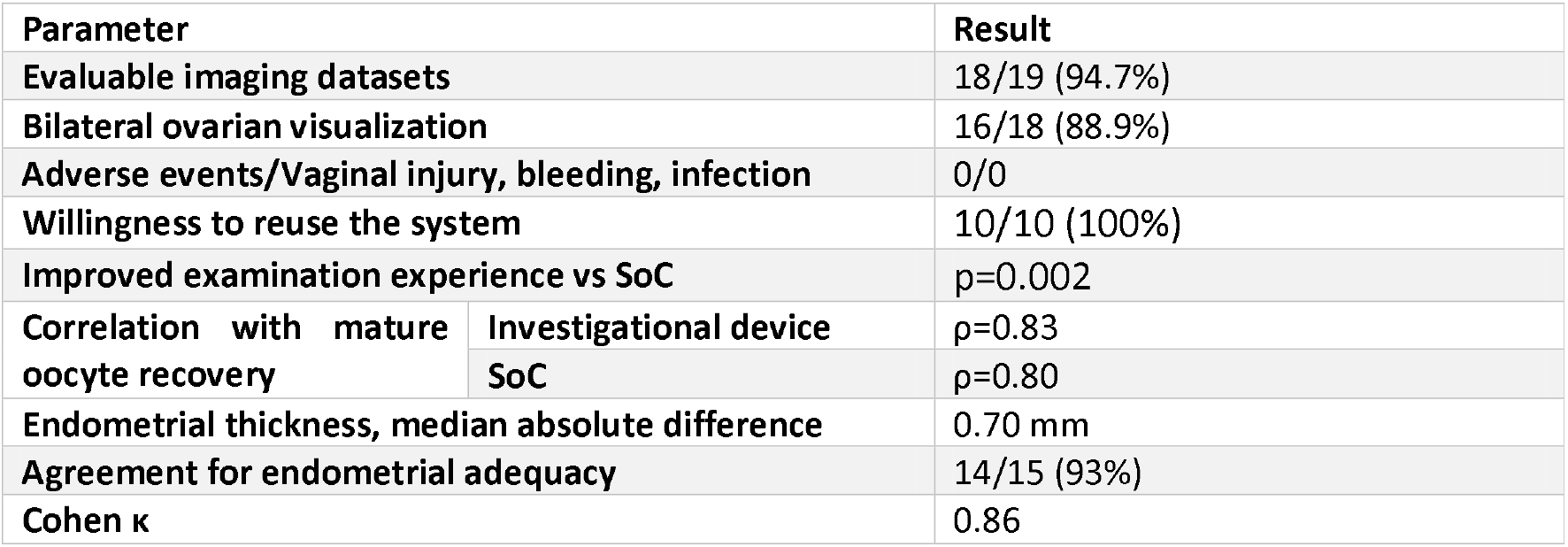
Key feasibility, clinical, and patient-reported outcomes.

| **Parameter** |  | **Result** |
|---|---|---|
| **Evaluable imaging datasets** |  | 18/19 (94.7%) |
| **Bilateral ovarian visualization** |  | 16/18 (88.9%) |
| **Adverse events/Vaginal injury, bleeding, infection** |  | 0/0 |
| **Willingness to reuse the system** |  | 10/10 (100%) |
| **Improved examination experience vs SoC** |  | p=0.002 |
| **Correlation with mature oocyte recovery** | **Investigational device** | ρ=0.83 |
|  | **SoC** | ρ=0.80 |
| **Endometrial thickness, median absolute difference** |  | 0.70 mm |
| **Agreement for endometrial adequacy** |  | 14/15 (93%) |
| **Cohen κ** |  | 0.86 |

**Figure 1.**
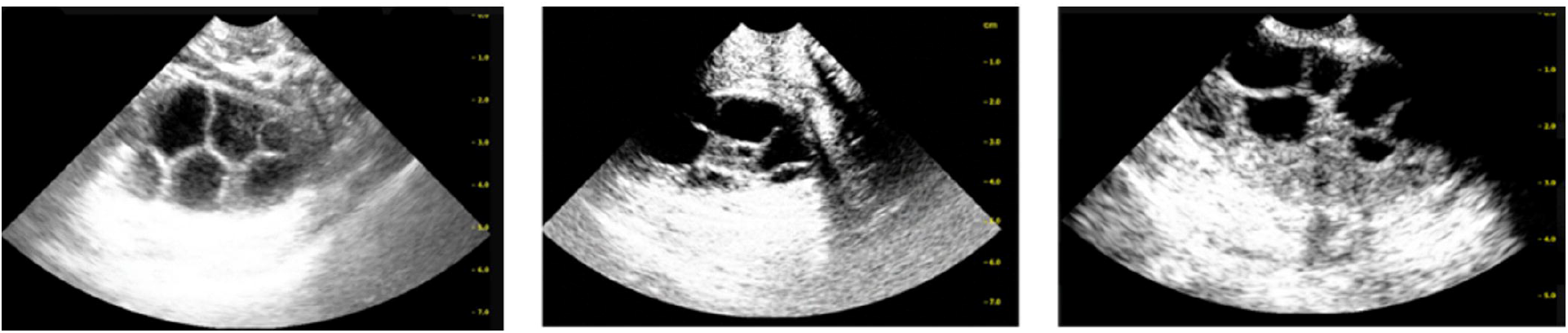
Typical ultrasound images during ovarian stimulation monitoring obtained by patients using the investigational prototype.

## Data Availability

All data produced in the present study are available upon reasonable request to the authors

## Supplementary Methods

### Investigational device and acquisition workflow

The investigational system consisted of a two-dimensional transvaginal ultrasound probe integrated with an external motorized rotational mechanism and acquisition control interface. Participants inserted and stabilized the probe themselves and received no formal sonographic training or real-time anatomical coaching.

During acquisition, the distal portion of the probe was rotated around its longitudinal axis over an approximately −180° to +180° range according to predefined angular steps. The rotational sequence was predetermined and was not modified according to the ultrasound images displayed during acquisition. In the prototype evaluated in this study, the external rotation mechanism was operated by a supervising healthcare professional; however, the professional did not navigate or reposition the probe according to real-time anatomy. Thus, professional involvement consisted of operation of the predefined rotational acquisition sequence rather than conventional sonographer-directed probe manipulation.

No automated image interpretation or treatment recommendation was generated by the investigational system, and investigational imaging was not used for clinical decision-making. The clinical study report similarly specifies that participants inserted and held the probe, external motor rotation was operated by the supervisor, and no real-time medical decisions were based on investigational imaging.

### Standard-of-care comparator

Each investigational examination was performed immediately after routine ovarian stimulation monitoring. Standard transvaginal ultrasound examinations were performed by two experienced sonographers using a Voluson S10 Expert ultrasound system (GE HealthCare, Chicago, IL, USA) with a two-dimensional transvaginal probe. One sonographer served as the primary operator throughout the study; the second had been trained according to the center’s standardized ovarian stimulation monitoring protocol. Both routinely performed ovarian stimulation monitoring in clinical practice. Standard ultrasound findings served as the clinical reference and were used for all treatment decisions.

### Preclinical evaluation and investigational device readiness

Before clinical use, the investigational prototype underwent documented technical, safety, and preclinical evaluation. Patient-contacting materials of the Vermon ultrasound transducer were evaluated according to ISO 10993-1, and electrical insulation testing included dielectric strength and leakage-current requirements. The Cephasonics Cicada ultrasound acquisition platform was operated with a medical-grade power supply tested to IEC 60601-1 and electrical isolation providing two means of patient protection (2 MOPP).

Preclinical evaluation of the integrated prototype included anthropomorphic pelvic phantom testing to assess image quality, depth penetration, measurement accuracy, and acquisition consistency, as well as CAD simulations evaluating probe positioning, insertion geometry, ergonomics, and field of view. Phantom follicular measurements were within ±5% of reference dimensions. A prospective risk analysis was completed before study initiation, and corresponding risk-control measures were incorporated into study procedures.

During the clinical investigation, the prototype was used adjunctively to standard-of-care ultrasound under study-staff supervision and was not used for clinical decision-making.

### Image review and analysis definitions

A scan attempt was defined as any investigational acquisition initiated during a study visit. An evaluable dataset was defined as an acquisition of sufficient quality to permit review of ovarian structures relevant to stimulation monitoring, with endometrial assessment performed when adequately visualized. Bilateral ovarian visualization and partial ovarian visualization were recorded separately.

Investigational image analysis included an initial blinded assessment followed by physician verification. Comparative follicular analyses were restricted to paired examinations and to ovaries visualized by both investigational and standard-of-care ultrasound. Follicles <9 mm were excluded from comparative analyses because they were not consistently individualized or reported during routine standard-of-care examinations. This approach was specified to improve comparability between modalities and reduce differences related to clinical reporting practices.

Counts of follicles ≥14 mm on the final pre-trigger examination were correlated with mature oocyte recovery. Endometrial analyses were restricted to examinations in which paired measurements were available. Statistical analyses were exploratory and unadjusted and were performed on paired datasets where applicable.

### Technical device deficiencies

Three non-serious prototype-related technical device deficiencies occurred during the clinical investigation.

Two examinations were associated with increased resistance of the probe rotation mechanism. Subsequent investigation determined that a progressive twisting of the internal cable bundle occurred during rotation. In both cases, rotation was completed with gentle manual assistance of the cable bundle, and the examinations were successfully completed. Both events occurred on the same study day before the common root cause had been fully identified. Root-cause analysis identified a device-setup procedural error rather than failure of the internal rotation mechanism. Corrective and preventive actions included inspection and functional verification of the device, retraining of study personnel, and addition of mandatory verification of the cable bundle to the pre-use checklist. No recurrence occurred after implementation of these measures.

A third event involved fracture of an externally mounted prototype rotation handle during transportation. Inspection confirmed that the internal rotation mechanism remained functional. The examination was completed by direct manual actuation of the rotation pulley according to the predefined degraded operating procedure. The damaged component was subsequently replaced, full device function was verified, and additional mechanical protection was added to the transport case. This event did not recur.

None of the three device deficiencies resulted in participant injury, additional clinical risk, examination termination, medical intervention, treatment interruption, or impact on clinical management. All three associated acquisitions remained evaluable.

### Endometrial visualization

Endometrial thickness measurements were available for 15 of 18 paired examinations. In three examinations, the endometrium was not adequately visualized with the investigational device because the probe settings and predefined acquisition sequence did not provide an appropriate imaging plane. Because the probe was not manually repositioned or adjusted in response to real-time anatomy during acquisition, an endometrial thickness measurement could not be obtained in these cases.

These cases reflect a technical limitation of the investigational prototype and fixed acquisition configuration evaluated in this study. Probe configuration and acquisition parameters are being refined during ongoing device development to improve consistent endometrial visualization.

## Notes

**Disclosure:** Tal Shavit reports no conflicts of interest. Pietro Bortoletto reports unpaid consulting activities for IMMA health outside the submitted work. Johanna Szychter is a stockholder of IMMA health. Yael Corcos reports consulting activities for IMMA health outside the submitted work. Sari Mendel reports no conflicts of interest. John Petrozza reports unpaid consulting activities for IMMA health outside the submitted work. Nadia Prisant is an employee, company officer, and stockholder of IMMA health.

**AI Disclosure Statement:** The authors declare that no artificial intelligence (AI)-assisted technologies were used to generate scientific content, interpret study findings, or draw scientific conclusions in this manuscript. AI-assisted tools were used solely for language editing and formatting support, with full human review and responsibility retained by the authors.

### Competing Interest Statement

Disclosure: Tal Shavit reports no conflicts of interest. Pietro Bortoletto reports unpaid consulting activities for IMMA health outside the submitted work. Johanna Szychter is a stockholder of IMMA health. Yael Corcos reports consulting activities for IMMA health outside the submitted work. Sari Mendel reports no conflicts of interest. John Petrozza reports unpaid consulting activities for IMMA health outside the submitted work. Nadia Prisant is an employee, company officer, and stockholder of IMMA health.
AI Disclosure Statement: The authors declare that no artificial intelligence (AI)-assisted technologies were used to generate scientific content, interpret study findings, or draw scientific conclusions in this manuscript. AI-assisted tools were used solely for language editing and formatting support, with full human review and responsibility retained by the authors.

### Author Declarations

Ethics committee/IRB of Assuta Medical Center gave ethical approval for this work

### Summary of Updates

This paper has been submitted in a research letter format following reviewers' appraisals.

## References

1. Gerris J, Delvigne A, Dhont N, Vandekerckhove F, Madoc B, Buyle M, et al. Self-operated endovaginal telemonitoring versus traditional monitoring of ovarian stimulation in assisted reproduction: an RCT. Hum Reprod. 2014;29:1941–8.

2. Shufaro Y, Cohen M, Wertheimer A, et al. Monitoring ovarian stimulation for assisted reproduction with patient self-scans using a home vaginal ultrasound device: a single-center interventional, prospective study. J Med Internet Res 2025;27.

3. Abbara A, Vuong LN, Ho VNA, Clarke SA, Jeffers L, Comninos AN, et al. Follicle size on day of trigger most likely to yield a mature oocyte. Front Endocrinol (Lausanne). 2018;9:193.

4. Forman RG, Robinson J, Yudkin P, Egan D, Reynolds K, Barlow DH. What is the true follicular diameter? An assessment of the reproducibility of transvaginal ultrasound monitoring in stimulated cycles. Fertil Steril 1991;56:989–92.

5. Wittmaack FM, Kreger DO, Blasco L, Tureck RW, Mastroianni L Jr, Lessey BA. Effect of follicular size on oocyte retrieval, fertilization, cleavage, and embryo quality in in vitro fertilization cycles: a 6-year data collection. Fertil Steril. 1994;62(6):1205–10.

